# Knowledge, Attitude, and Practice of Vasoactive Agents Infusions : Development and Psychometric Properties of a Questionnaire with Chinese Clinical Nurses

**DOI:** 10.1101/2024.10.01.24314752

**Authors:** Yanfang Huang, Yi Chen, Yaping Fang, Junjie Mou, Shiqin Li, Xianying Lei, Yuxin Li, Zhongping Ai

## Abstract

**Background:** Inconsistencies with guidelines or standards regarding nurses’ practice of vasoactive agent infusion have been documented. Adequate knowledge and positive attitudes are critical for compliance. However, there are currently no validated tools specifically designed to measure the knowledge and attitudes related to vasoactive agent infusions among nurses.

**Objective:** The aim of this study was to develop and test the validity and reliability of the Chinese mainland version of Knowledge, Attitude, and Practice of Vasoactive Agents Infusions Questionnaire among nurses.

**Methods:** The initial questionnaire items were developed through a comprehensive literature review, expert consultation, and pilot study. From February to June 2024, cross-sectional data were collected using convenience sampling from 538 nurses across 9 hospitals in Sichuan Province, China. The reliability and validity of the scale were evaluated through internal consistency reliability, inter-rater reliability, exploratory factor analysis, and confirmatory factor analysis.

**Results:** The final version of the questionnaire included 33 items across 3 dimensions, explaining 78.04% of the variance, with item loadings ranging from 0.56 to 0.89. The content validity index ranged from 0.91 to 1.00, and the scale-level content validity index was 0.98. The overall Cronbach’s α for the questionnaire was 0.96, with Cronbach’s α for each dimension ranging from 0.96 to 0.98. The test-retest reliability for the entire questionnaire was 0.90, and for each dimension, it ranged from 0.90 to 0.94 (p < 0.05). The confirmatory factor analysis demonstrated acceptable fit indices for the three-dimensional model: χ²/(df) = 1.135, p = 0.200, RMSEA = 0.024, CFI = 0.986, TLI = 0.985, GFI = 0.884, and NFI = 0.895.

**Conclusion:** The Knowledge, Attitude, and Practice of Vasoactive Agents Infusions Questionnaire demonstrates good reliability and validity, making it a reliable measurement tool for assessing nurses’ attitudes and knowledge related to vasoactive agents’ infusions. This version will facilitate further research and advancements in this specific field of study.

## 1. Introduction

Vasoactive agents are a class of drugs that regulate vascular tone, diameter, and blood flow. By acting on vascular smooth muscle and endothelial cells, they modulate vasoconstriction or vasodilation, thus influencing blood pressure and organ perfusion[1]. Between 10% and 54% of patients admitted to intensive care units (ICUs) have been reported to receive vasoactive agents for conditions such as sepsis, heart failure, and organ failure. Moreover, factors such as the dose and type of vasoactive drug used are associated with patient mortality[2-4]. Nurses were entrusted with all facets of medication management, encompassing infusion preparation, initiation, administration, titration, weaning, and documentation. Due to the complexity of vasoactive agents, nurses face significant challenges in making dynamic clinical decision by nurses in administering vasoactive medications in patients [5]. A latest observational study found that in Australia, nurses consistently maintain patients’ Mean Arterial Pressure (MAP) above target levels to prevent hypotension, frequently overlooking the potential adverse effects of high doses of vasoactive agents[6]. This poses a serious threat to patient safety, particularly for those with hemodynamic instability.

In China, a rising corpus of research on vasoactive agents’ administration may be attributed to the expansion of advanced practice nursing (APN) and specialized graduate training programs in nursing science. Early study demonstrated the feasibility of peripheral infusion of vasoactive agents[7] and summarized evidence for their safe management[8]. To give clinical practitioners methodical and scientific criteria, the Chinese Nursing Association established a group standard for “Nursing of Vasoactive Agent Infusion”. However, when it comes to nursing quality and treatment results, there is a lot of variation in nurses’ adherence to this guideline in clinical practice[9]. Significant challenges emerge when integrating evidence and guidelines into clinical nursing routine[10].

Inadequate implementation of vasoactive agents’ infusion protocols by clinical nurses can be attributed to a number of reasons. Despite the increasing emphasis on the safety administration of vasoactive drugs by nursing professionals in recent years, there remains limited insight into the level of knowledge and attitudes employed by clinical nurses[11]. To achieve optimal outcomes, nursing administrators need to comprehensively understand the knowledge, attitudes, and practices of clinical nurses concerning vasoactive agent infusions. Hence, this study aimed to develop the Knowledge, Attitude, and Practice of Vasoactive Agents Infusions Questionnaire (KAPV-AIQ) for nurses. This tool will help identify weaknesses in practice through questionnaire assessments, guide the development of improvement measures. Consequently, it is expected to improve overall nursing quality, ensure patient safety and treatment efficacy, and aid in formulating more detailed and actionable training programs.

## 2. Materials and Methods

The study consisted of the following phases:

### 2.1 Phases1: Instrument Development

#### 2.1.1 Specify dimensional structure

Firstly, we rigorously applied the Knowledge, Attitude, and Practice (KAP) theory[12] as a guiding framework to identify the dimensions of our questionnaire. Subsequently, our expert panel reached a consensus on the conceptualization of these dimensions, drawing from literature review and considering the specific context of clinical nursing care in China. This consensus was achieved after two rounds of thorough discussions. Nurses’ Knowledge of Vasoactive Agent Infusion refers to the comprehensive understanding and awareness that nurses possess regarding the administration of vasoactive medications. This includes their familiarity with the pharmacological properties, indications, contraindications, dosage calculations, infusion protocols, monitoring requirements, potential adverse effects, and emergency management associated with these potent medications. Nurses’ Attitude Towards Vasoactive Agent Infusion encompasses their beliefs, perceptions, and feelings about the administration and management of vasoactive medications. This includes their confidence in handling these potent drugs, their commitment to adhering to safety protocols, their sense of responsibility in monitoring and managing patients receiving such infusions, and their overall perspective on the importance and impact of vasoactive agent administration on patient outcomes. Nurses’ Practice of Vasoactive Agent Infusion refers to the actions and procedures that nurses perform in the administration of vasoactive medications. This involves the correct preparation, dosage calculation, and administration of these drugs, continuous patient monitoring, prompt identification and management of adverse reactions, adherence to clinical protocols and guidelines, and accurate documentation of the infusion process.

#### 2.1.2 Item generation

A systematic literature review was undertaken to gather comprehensive information, using key terms including “vasoactive drugs” “vasopressors” “inotropes” “infusion care” “intravenous infusion” “medication administration” “safety management” “patient safety” “risk management” and “adverse effects”. Databases searched included PubMed, Cochrane Library, Embase, and Scopus. The search timeframe extended from the inception of each database to December 2023. Boolean operators (AND, OR NOT) were employed to refine and combine search terms. After two independent team members completed the initial screening, the final selection of target literature was decided through group consensus. This process resulted in the inclusion of two guidelines[13,14], two expert consensuses[15-16]; one evidence summaries[11]; and the Health of the People’s Republic of China industry standards (WS/T 433—2023) and the Chinese Nursing Association Group Standard (T/CNAS 22─2021) for synthesis. Using these sources, we developed a preliminary questionnaire encompassing three dimensions (knowledge, attitude, practice) with a total of 43 items.

#### 2.1.3 Item screening

In this study, items were screened using correlation coefficient analysis factor analysis and Cronbach’s α coefficient analysis. The correlation coefficient method was utilized to analyze the relationship between each item and its respective dimension. Items were retained if they demonstrated a high correlation with their own dimension and a low correlation with other dimensions. Additionally, an item was preserved if its correlation coefficient with the total scale score (r) was greater than 0.4 and statistically significant (P<0.05). Subsequently, factor analysis was conducted to extract factor loadings for each item. The criteria for item deletion were as follows: factor loading ≤ 0.5, communality ≤ 0.2, and each dimension required a minimum of three items [17]. During the item selection process, internal consistency of the questionnaire was also considered. Specifically, if the Cronbach’s alpha coefficient increased significantly when an item was deleted, it indicated that the item might reduce the internal consistency of the dimension, and thus, the item was considered for deletion. Finally, 239 valid questionnaires were used to factor analysis for screening items.

Correlation analysis indicated that in the Knowledge dimension, the items “K13. Are you aware that central venous access is preferred for vasoactive agent infusion, with peripheral large vein access as an option in emergencies?” and “K9. Are you aware of the key aspects of patient condition that should be closely monitored during vasoactive agent use?” had correlation coefficients of 0.215 and 0.123, respectively, with the total questionnaire score. In the Behavior dimension, the item “P29. When adjusting the dose of vasoactive agents, I accurately and thoroughly record the drug name, dosage, concentration, infusion rate, change time, blood pressure, heart rate, and rhythm” had a correlation coefficient of 0.225 with the total questionnaire score. These items were therefore excluded. The remaining 40 items had correlation coefficients ranging from 0.557 to 0.904 with the total questionnaire score, all exceeding 0.4 and showing statistical significance (P < 0.001).

Subsequent factor analysis led to the removal of items with factor loadings below 0.4, including: “K10. Are you aware that an infusion pump should be used for administering vasoactive agents?” “K18. Are you aware of the pH of commonly used vasoactive agents?” “K19. Are you aware of the half-life of commonly used vasoactive agents?” “K20. Are you aware of the onset time of commonly used vasoactive agents?” “K21. Are you aware of the metabolic pathways of commonly used vasoactive agents?” “K22. Are you aware of the initial dose for commonly used vasoactive agents?” and “K23. Are you aware of the routine adjustment doses for commonly used vasoactive agents?”

#### 2.1.4 Pilot survey

In February 2024, a pilot survey was conducted using convenience sampling to select nurses from a tertiary teaching hospital in Sichuan Province. The survey was administered online. Inclusion criteria for participants were possession of a valid nursing license, formal employment by the hospital, engaged in frontline clinical nursing work, and voluntary participation in the survey. Exclusion criteria included nurses on sick leave, maternity leave, or away for training purposes. Participants evaluated the questionnaire using a Likert 5-point scale, assessing several key aspects: the rationality of the questionnaire design, the clarity and comprehensibility of the text, feasibility, willingness to self-assess using the questionnaire, comfort level while answering, and willingness to discuss the questionnaire with researchers. Issues and suggestions raised by participants were also collected and documented.

A total of 60 valid questionnaires were returned. All participants found the questionnaire design rational, the text clear and comprehensible, and the questionnaire feasible. They expressed willingness to use the questionnaire for self-assessment, felt comfortable during the answering process, and were willing to discuss the questionnaire content with other researchers. The average time to complete the questionnaire was 218 seconds.

#### 2.1.5 Content Validity Evaluation

In this study, we engaged 11 experts, including 2 critical care medicine specialists, 5 critical care nursing specialists, 2 nursing quality management experts, 1 nursing research expert, and 1 clinical pharmacy expert. These experts evaluated the content validity, linguistic clarity, and clinical relevance of the questionnaire items based on their clinical experience and professional expertise and provided recommendations for revisions.

Content validity was assessed using a 4-point Likert scale, where scores ranged from 1 (“not relevant”) to 4 (“highly relevant”). The item-level content validity index (I-CVI) was calculated as the proportion of experts rating an item as 3 or 4 out of the total number of experts. The scale-level content validity index/average (S-CVI/Ave) was determined as the percentage of items rated 3 or 4 by all experts, indicating overall consensus. Items with an I-CVI of 0.78 or higher from at least three experts and an S-CVI/Ave of 0.90 or higher were considered to demonstrate acceptable content validity[18].

We collected 11 valid responses. All experts held at least an associate senior professional title and had a master’s degree or higher, with an average of 26.00 years of professional experience (SD = 8.83 years). To enhance clarity, the experts recommended the following revisions: changing “adverse reactions of vasoactive agents” to “common adverse reactions of vasoactive agents”; revising “monitoring blood pressure, heart rate, heart rhythm, etc.” to “monitoring vital signs”; and updating “selection of infusion routes for vasoactive agents” to “principles for selecting infusion routes for vasoactive agents.” Regarding item 7, “Do you know the specifications of vasoactive agents?” one expert suggested its removal due to limited relevance. However, after review and factor analysis (with factor loading > 0.5, see Table 1), the item was retained.

**Table 1.** Exploratory factor analysis structure matrix of Knowledge, Attitude, and Practice of Vasoactive Agents Infusions Questionnaire (N=239)

| Items | Factors |  |  |
| --- | --- | --- | --- |
|  | Knowledge | Practice | Attitude |
| K1.Definition of vasoactive agent | **.827** | .177 | .194 |
| K2.Classification of vasoactive agent | **.865** | .123 | .173 |
| K3.Common adverse reactions of vasoactive agent | **.856** | .107 | .190 |
| K4.Principles of selecting infusion routes for vasoactive agent | **.855** | .246 | .167 |
| K5.Requirements for infusing vasoactive agent | **.849** | .226 | .112 |
| K6.Proper configuration of vasoactive agent | **.868** | .229 | .076 |
| K7.Specifications of commonly used vasoactive agent | **.898** | .169 | .109 |
| K8.Precautions during the infusion process of commonly used vasoactive agent | **.895** | .231 | .108 |
| K9.Use of pump for infusing vasoactive agent | **.836** | .314 | .128 |
| K10.Correct use of infusion pumps | **.829** | .321 | .091 |
| K11.Common alarms and management of infusion pumps | **.789** | .343 | .113 |
| K12.Labeling requirements for infusion ports when simultaneously infusing multiple vasoactive agent | **.771** | .361 | .122 |
| K13.Conditions appropriate for using the dual-pump method for continuously infusing vasoactive agent | **.836** | .209 | .102 |
| K14.Monitoring frequency of vital signs during initial use, dose adjustment, and stabilization of vasoactive agent | **.832** | .287 | .088 |
| K15.Procedure for stopping vasoactive drug infusion | **.779** | .213 | .157 |
| A4.Recognizing the vital importance of clinical nurses mastering vasoactive agent care knowledge | .194 | .299 | **.845** |
| A2.Emphasizing the vital role of dynamic assessment during vasoactive agent infusion | .195 | .300 | **.872** |
| A3.Commitment to integrating standards of vasoactive agent infusion into clinical practice | .188 | .337 | **.920** |
| A5.Highlighting the importance of regular specialized training on vasoactive agent infusion | .151 | .326 | **.861** |
| A1.The crucial role of guidelines for vasoactive agent infusion in clinical practice | .163 | .332 | **.854** |
| P4.Vigilant monitoring of infusion pathway and surrounding skin conditions is conducted throughout. | .290 | **.678** | .340 |
| P10.Continuous observation of infusion rate and remaining drug volume is maintained. | .296 | **.751** | .321 |
| P5.Each syringe used during infusion is labeled with patient details, drug specifics, and preparation information. | .196 | **.787** | .200 |
| P12.Prompt replacement of the infusion line is ensured upon reflux detection. | .345 | **.700** | .329 |
| P6.Both ends of the infusion line are clearly labeled during setup. | .355 | **.560** | .172 |
| P13.Preparations for continued infusion are made as drug completion nears, ensuring seamless continuation. | .244 | **.824** | .216 |
| P7.Syringe and infusion tubing are replaced when changing medications. | .284 | **.746** | .126 |
| P11.Collaboration with physicians ensures alignment with therapeutic goals during infusion, with ongoing adjustments and monitoring of vital signs. | .231 | **.858** | .159 |
| P1.Prior to vasoactive agent administration, a dual-person check ensures prescription accuracy regarding dose, method, and infusion rate, with immediate physician clarification if needed. | .219 | **.852** | .227 |
| P2.Assessment includes verifying vascular access, infusion pump functionality, and battery status before drug administration. | .244 | **.797** | .212 |
| P8.During connection of infusion lines, ensuring medication flows correctly to the interface is prioritized. | .184 | **.846** | .154 |
| P3.Precise preparation of drug concentration and infusion rate is conducted according to prescription guidelines before starting infusion. | .149 | **.860** | .219 |
| P9.Ongoing assessment of patient vital signs, peripheral circulation, and urine output is integral during drug administration. | .225 | **.857** | .183 |
| Eigenvalues | 18.19 | 5.14 | 2.42 |
| % of Variance Explained | 35.16 | 28.49 | 14.38 |
| Cumulative % of Variance Explained | 35.16 | 63.65 | 78.04 |

### 2.2 Phases2: Test the Psychometric Properties of the Questionnaire

#### 2.2.1 Design, setting and sample

A cross-sectional study was conducted from February 2024 to June 2024. Convenience sampling was used to select clinical nurses from 6 tertiary hospitals and 3 secondary hospitals in Sichuan Province. Eligibility and exclusion criteria were consistent with the pilot survey. The instruments comprised a general information questionnaire (covering age, gender, education, title, and years of clinical experience et al.) and the KAP- VAIQ. To assess test-retest reliability, 20 respondents were conveniently selected for a follow-up survey two weeks after the initial data collection.

The estimated minimum sample size required for factor analysis was 330 to 660 participants, following the recommended item-to-response ratio is between 10:1 and 20:1[19]. With the KAP-VAIQ containing 33 items, excluding the 60 nurses involved in the acceptability evaluation and the 20 nurses used for test-retest reliability.

#### 2.2.2 Procedures

From February 6th, 2024, online recruitment advertisements and questionnaire link were disseminated via WeChat and the WenJuanXing platform to invite voluntary participation from nurses at selected hospitals. The advertisements included study details and contact information. Participants were required to review the study introduction and consent form before accessing the questionnaire. Only after providing informed consent could, they proceed with completing the questionnaire. Data were downloaded from the platform after survey completion, ensuring no duplicate responses due to the platform’s unique identifier system.

#### 2.2.3 Internal Reliability and Test-Retest Stability

Cronbach’s alpha coefficient was used to express the internal consistency of KAPV- AIQ. The internal consistency coefficient greater than 0.7 indicates high reliability of the scale[20]. The test-retest reliability and internal consistency were assessed for reliability. The intraclass correlation coefficient (ICC) was used to express the retest reliability, which is equal to the individual variability divided by the total variability. Its range of values is between 0 and 1. In general, a reliability coefficient below 0.4 indicates poor reliability, and above 0.75 means good reliability[21].

#### 2.2.3 Exploratory Factor Analysis and Confirmatory Factor Analysis

Exploratory factor analysis (EFA) and was Confirmatory factor analysis (CFA) were used to examine the construct validity. Kaiser-Meyer-Olkin (KMO) and Bartlett’s test of sphericity were considered to check the adequacy and suitability of the sample data for factor analysis. Principle Component Methods (PC) and Varimax with Kaiser Normalization were used to do the factor extraction to identify meaningful items. KMO value should be more than 0.5 and Bartlett’s test had a probability of <0.05 which means that correlational matrix could be factorable[22]. The criterion for factor extraction is to retain factors with eigenvalues greater than 1. The model fit of the SEM was reflected by the fit indices including the model chi-square (χ^2^), Goodness of Fit Index, GFI, Root Mean Square Residual RMR, Root Mean Square Error of Approximation, RMSEA, Comparative Fit Index, CFI, Goodness of Fit Index, GFI, Normed Fit Index, NFI, Tucker-Lewis Coefficient, TLI[23].

### 2.3 Ethical Consideration

Ethical approval was obtained from Southwest Medical University Hospital, China, with ethical approval codes KY2024064. The informed consent will be posted first on the “Wenjuanxing” web application. Only if they agree will the questionnaires be displayed for them to complete. If they do not agree, they will be directed to “exit” the platform and will receive a “thank you” message. All participants have the right to refuse or withdraw from the study at any time without penalty. All information gathered for this study stored on a password-protected computer in the form of electronic files and kept confidential from those who do not have the right to know.

### 2.4 Data Analysis

After the original data is downloaded from “Wenjuanxing” platform, it is sorted and imported into SPSS 28.0 through Excel. The demographic data of the participants, the reliability, and the EFA was described and analyzed using SPSS. The CFA was used the Analysis of Moment Structure (AMOS). In a 1:1 ratio, the initial 239 samples underwent EFA, and the subsequent 239 samples underwent CFA.

## 3. Results

### 3.1 The Final version of Questionnaire

The final version of KAP-VAICQ consists of three dimensions: Knowledge with 15 items, Attitudes with 5 items, and Practices with 13 items. All items use a 5-point Likert scale, with the Knowledge dimension ranging from “Not at all familiar” to “Very familiar”. The Attitudes dimension ranging from “Strongly disagree” to “Strongly agree” and the Practices dimension ranging from “Never” to “Always”. Scores are assigned from 1 to 5 for each item, with the total questionnaire score ranging from 33 to 165 points. A higher score indicates a greater level of knowledge, attitudes, and practices related to vasoactive agent infusion nursing among the nurses.

### 3.2 Content validity

The item-level Content Validity Index (I-CVI) for the questionnaire ranged from 0.91 to 1.00, while the scale-level Content Validity Index/average (S-CVI/Ave) was 0.98.

### 3.3 Participants’ characteristics

A total of 535 nurses participated in the psychometric assessment. Of these, 478 valid questionnaires were collected, yielding a response rate of 89.34%. The majority of participants were women (83.24%). The mean age of the participants was 32.86 years (SD = 7.18) with a median age of 32 years. A significant portion of respondents (81.63%) held a bachelor’s degree or higher. All participants were registered nurses, comprising 425 (88.91%) from tertiary hospitals and 53 (11.08%) from secondary hospitals. Specifically, there were 52 (10.87%) internal medicine nurses, 142 (29.70%) surgical nurses, 189 (39.53%) critical care nurses, 30 (6.27%) operating room/anesthesia nurses, 21 (4.39%) pediatric nurses, and 16 (3.34%) others. The distribution of professional titles included 253 (52.92%) with junior professional titles, 193 (40.37%) with intermediate professional titles, and 32 (6.69%) with senior professional titles or above. The average clinical nursing experience was 8.32 years (SD = 5.07)

### 3.4 Reliability

The Cronbach’s α coefficient for the overall questionnaire was 0.96, with Cronbach’s α coefficients for the individual dimensions ranging from 0.96 to 0.98. The intraclass correlation coefficient for test-retest reliability of the questionnaire was 0.90, and the intraclass correlation coefficients for the dimensions ranged from 0.90 to 0.94 (P < 0.05).

### 3.5 Exploratory factor analysis

EFA was conducted on 239 sample data. The KMO measure of sampling adequacy was 0.945, and Bartlett’s test of sphericity yielded a χ² value of 10,422.37 with a p-value of 0.00. Using principal component analysis and applying the criterion of eigenvalues greater than 1, 3 common factors were extracted, accounting for 78.04% of the cumulative variance. The factors’ loadings were computed using varimax rotation, revealing that all item loadings were greater than 0.50, with factor loadings ranging from 0.56 to 0.89 on their respective common factors **(see Table 1)**

### 3.6 Confirmatory factor analysis

The model yields the acceptable indices: χ^2^ /df=1.135, p=0.020, Goodness of Fit Index, GFI=0.884, Root Mean Square Residual RMR=0.030, Root Mean Square Error of Approximation, RMSEA=0.024, Comparative Fit Index, CFI=0.986, Goodness of Fit Index, GFI=0.884, Normed Fit Index, NFI=0.895, Tucker-Lewis Coefficient, TLI=0.985.

**Figure 1.**
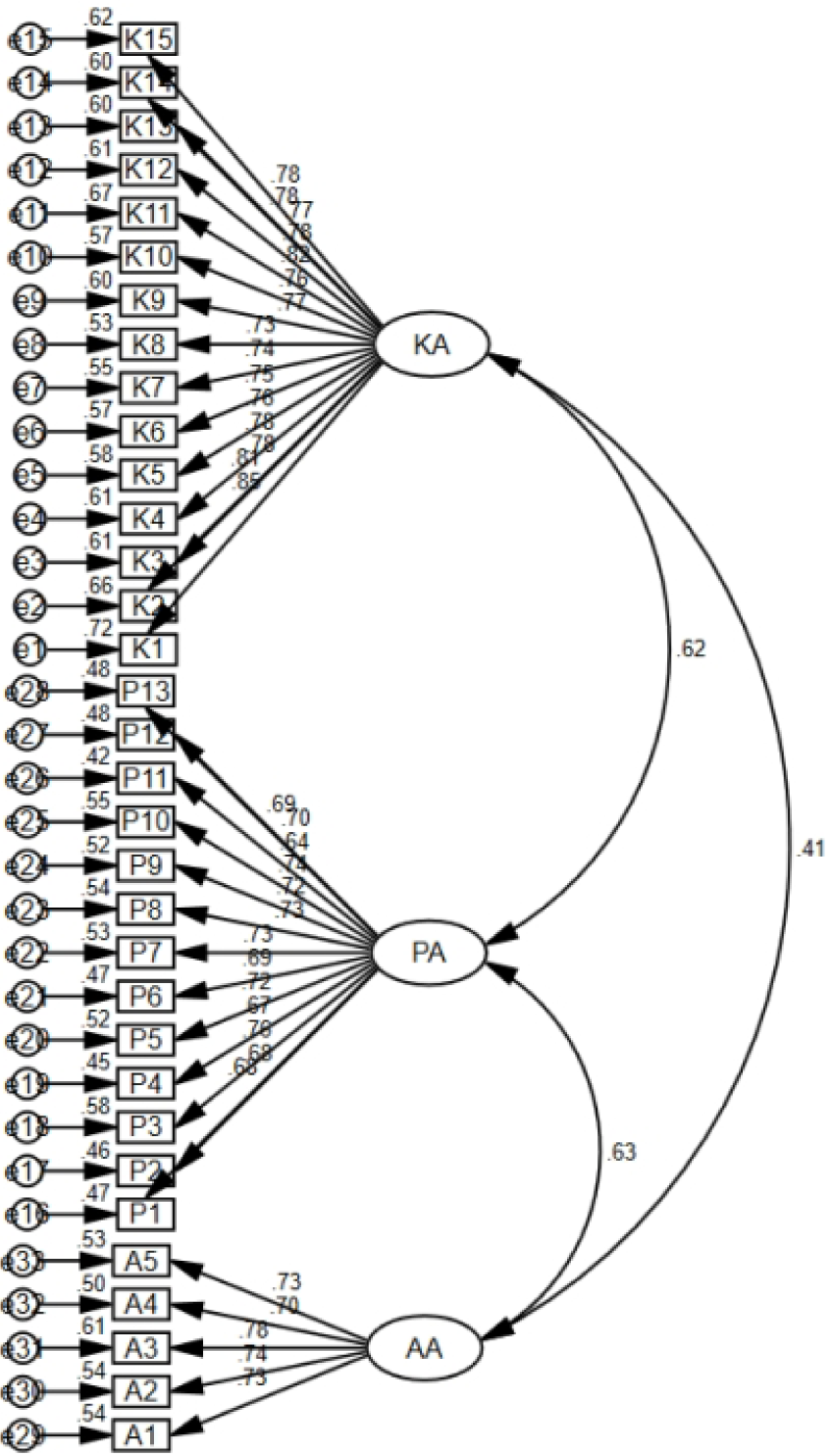
Confirmatory factor analysis of Knowledge, Attitude, and Practice of Vasoactive Agents Infusions Questionnaire ( N=239)

## 4. Discussion

In this study, we developed and tested a preliminary Chinese questionnaire to understand the knowledge, attitudes, and practices of clinical nurses concerning vasoactive agent infusions. In time as we knew, this had never been conducted previously. Most of the existing measurements of nurses’ knowledge and practice of vasoactive agents were developed based on empirical researchers, and psychometric indicators were not reported, affecting the accurate assessment objectively as well as limitations of external validation and outreach[24,25]. However, it is crucial to design educational curricula for the administration of vasoactive drugs based on clinical nurses’ quantitative and measurable levels of knowledge and practice[26].

The final version of KAP-VAICQ, three self-assessments dimensions of knowledge with 15 items, attitude with 5 items, and practice with 13 items, were designed at the individual nurse level, which qualities were consistency with the Theory of Knowledge, Attitude, and Practice (KAP). The KAP theory hypothesis states that adopting practices and forming attitudes and knowledge are the three sequential processes that lead to human behavioral change[27]. In this study context, it is vital to improve the standardization practices of nurses in the administration of vasoactive agents to improve the safety of patients’ medication and the quality of care. Hence, this scale enables nursing administrators and researchers to efficiently assess nurses’ knowledge, attitudes and practices regarding vasoactive agent infusion. It also helps identify potential facilitators and barriers in the nursing process. Based on the assessment results, nursing administrators can enhance clinical nurses’ adherence to the standardized management of vasoactive agents by implementing targeted initiatives such as increasing relevant training, optimizing workflows, and ensuring resource availability. Due to the complexity of vasoactive agent management, this questionnaire remains to be further explored in terms of extravasation. In addition, studies have shown that factors were not involved in this questionnaire, such as intelligent technology [28,29] and organizational safety climate [30] may also influence the level of nurses in the management of vasoactive agent infusion.

In this study, Cronbach’s alpha coefficient and retest reliability method were used to assess the internal and external reliability of the questionnaire. The study results demonstrated strong internal consistency within questionnaire. Retest reliability analysis indicated good repeatability, and content validity assessment confirmed a high alignment between the measured content and the intended objectives. Besides, to assess the consistency and stability of the questionnaire framework, two different sets of data were used for EFA and CFA. In EFA, the Principal Component (PC) methods and Varimax with Kaiser Normalization were chosen as the preliminary solution [30]. After analysis, we obtained 3 factors that explained 35.16%, 28.49%, 14.38% respectively of the variance, and the accumulated percentage of explained variance was 78.04%. According to the evaluation criteria of CFA indicator [31], the results of yields the acceptable indices, indicating a good fit to the theoretical structure of KAP.

## limitations

There are some limitations in this study. Since this study used convenience sampling and the web-based questionnaire management platform showed that the IP addresses of all respondents were located in one province in western China. In addition, for the validity examination of this scale, there was no criterion validity test applied. The criterion validity of KAP-VAIQ may be tested in future studies.

## Conclusions

The Knowledge, Attitude, and Practice of Vasoactive Agents Infusions Questionnaire has good reliability and validity and can be used as a reliable measurement tool when evaluating nurses’ attitudes and knowledge to vasoactive agents’ infusions.

## Data Availability

All relevant data are within the manuscript and its Supporting Information files.

## Acknowledgments

The authors would like to express their gratitude to all hospital administrators for their support during the data collection process. The authors would also like to thank all the participants in this study.

